# Prevalence of HIV1 Infection and Its Associated Factors among Exposed Infants at Shegaw Motta General Hospital, Ethiopia

**DOI:** 10.1101/2024.09.01.24312902

**Authors:** Destaw Kebede Nigusie, Fantahun Getaneh Damitew, Kirubel Endalamaw Melsew, Girma Zerefaw, Abebe Feneta Nigusie

**Affiliations:** Department of Diagnostic Medical Laboratory Science at Shegaw Motta General Hospital, East Gojjam, Motta Town, Ethiopia; Department of Bacteriology and Mycology Reference Laboratory, Amhara Public Health Institute (APHI)-Debre Markos Branch, Debre Markos Town, Ethiopia; Department of Molecular Biology and Virology Reference Laboratory, Amhara Public Health Institute (APHI), Bahir Dar Town, Ethiopia; Department of Medical Laboratory science, College or Medicine and Health science, Debre Markos University, Debre Markos Town, Ethiopia

**Keywords:** HIV infection, associated factors, exposed infants, Ethiopia

## Abstract

**Background:** Human immunodeficiency virus (HIV)/acquired immunodeficiency syndrome is a leading cause of death and disease burden. Following this, vertical transmission is the main source of HIV infection on children globally. Morbidity and mortality among HIV-exposed infants are still the main health challenges in Ethiopia. Therefore, the aim of this study was to determine the prevalence of HIV1infection and its associated factors among exposed infants at Shegaw Motta General Hospital, Ethiopia.

**Methods:** Hospital-based cross-sectional study was conducted on exposed infants at Shegaw Motta General Hospital from September 1, 2022 to July 30, 2023. The consecutive convenience sampling technique was used to select study participants. Whole blood sample was collected from mothers and infants. Laboratory tests like early infant diagnosis, cluster of differentiation 4 counts and viral load were performed using standard operating procedure. Then, the data were entered into EpiData version 3.1 and analyzed by SPSS version 20. Finally, bivariable and multivariable logistic regressions were carried out to identify factors significantly associated (*P*<0.05).

**Results:** Out of 155 infants, about 79(50.9%) infants were females and87(56.1%) was urban resident. Furthermore, majority of infants were born from mothers who could not able to write and read 88(56.8%) and maternal ages range from 25-34years were 138(89.0%). The overall prevalence of HIV1 infection among exposed infants was6(3.87%) with (95%CI: 2.9-8.2). Pregnant women had not antennal care (AOR=7.281, *P* = 0.001), home delivery (AOR= 3.239, *P*=0.001), maternal not received antiretroviral prophylaxis (AOR = 9.213, *P*= 0.001), infants not intake nevirapine prophylaxis (AOR=2.560, *P*= 0.007) and maternal high viral load (AOR= 5.120, *P*= 0.004) were the factors associated with HIV infection among exposed infants.

**Conclusion:** The HIV1 infection among exposed infants was still high (3.87%). Pregnant women had not antenatal care follow up, home delivery, maternal high viral load, and not receiving antiretroviral prophylaxis, infant not intake nevirapine prophylaxis increases the risk of HIV1 infection. Therefore, health facilities should strictly strengthen the PMTCT service by providing maternal antiretroviral prophylaxis, infant nevirapine prophylaxis, promoting antenatal care service, early screening maternal viral load and scale up skilled delivery to eliminate HIV infection among exposed infants.

## Introduction

Human immunodeficiency virus (HIV)/acquired immunodeficiency syndrome (AIDS) is a leading cause of death and disease burden [1]. Following this, vertical transmission is the main source of HIV infection in children with an estimated 2000 vertically-acquired HIV infections occurring daily in global and mostly in Eastern Europe and Central Asia [2]. However, about 330,000 children were infected with HIV in 2011 globally, with over 90% of these infections through mother-to-child transmission was illustrating in sub-Sahara Africa [3].

Ethiopia is one of these priority countries where every 3 children born to women living with HIV still gets infected with HIV [4, 5]. So, infants acquire infection with HIV-1 through mother-to-child transmission (MTCT) of the virus [6]. MTCT of HIV-1 can occur through intrauterine (IU), at the time of labor and delivery or intra-partum, and postpartum mainly through breastfeeding [7,8].In addition to prenatal antiretroviral therapies, public health strategies such as prevention of maternal nipple lesions, mastitis and infant thrush; reduction of breastfeeding duration by all HIV-1-infected mothers; absolute avoidance of breastfeeding by those at high risk, and prevention of HIV-1 transmission to breastfeeding mothers should be addressed [9].

HIV-1 infection in breastfed children born to infected mothers is associated with the presence of integrated viral deoxy-ribose nucleic acid (DNA) in the mothers’ milk cells. IgM and IgA anti-HIV-1 in breastmilk may protect against postnatal transmission of the virus [10]. However, international guidance currently states that when replacement feeding is acceptable, feasible, affordable, sustainable and safe, the avoidance of all breastfeeding by HIV-infected mothers is recommended [11].

Most of the deaths in children with HIV could have been avoided through early infant diagnosis (EID) and provision of effective care and treatment. Interventions like the use of antiretroviral (ART) drugs by infected pregnant women, safe delivery practices and safe infant feeding have helped to reduce the risk of transmission to infants by 40% to 50% [12]. Because of passively transferred maternal HIV-1 antibodies, which may be present in the child’s bloodstream until 18 months of age, antibody tests are not reliable for diagnosing children less than 18 months of age [12, 13].Instead, polymerase chain reaction (PCR) such as EID provides a feasible method to assess prevention of mother-to child transmission (PMTCT) programs and early identify HIV-infected infants [14] the reason why its high sensitivity and specificity, PCR has been widely used for diagnosis of HIV amongst exposed infants as well as identification of infection from birth [15] Ethiopia is among the top ten countries in the world with the highest burden of HIV1 infections among children [16] with the average number of MTCT of HIV in Ethiopia was 18% [17]. So that, Child morbidity and mortality among HIV-exposed infants are still the main health challenges in Ethiopia [18]. However, there need a further study beyond HIV 1 and its associated factors among infants born to HIV positive women. Therefore, the aim of this study was to determine the prevalence of HIV 1and its associated factors among infants born from HIV positive mothers at Shegaw Motta General Hospital, Ethiopia.

## Materials and Method

### Study design, area and period

Hospital based cross sectional study was conducted at Shegaw Motta General Hospital (SMGH) which is found in East Gojjam zone, Amhara regional state. Shegaw Motta General Hospital is found in Amhara region, which is located at a distance of 120 Km from Bahir Dar and 370 Km away from the capital city of the Ethiopia, Addis Ababa. SMGH has provided service to more than 1.5 million total populations. Whenever, this study was conducted from September 1, 2022 to July 30, 2023.

### Study Population and participants

All infants (ages < 12months) born from HIV positive mothers were the study population. While, all HIV exposed infants attending PMTCT clinic in Shegaw Motta General Hospital and providing a blood sample during the study period were study participants.

### Sample size and sampling technique

The sample size was calculated by using single population proportion formula by taking the prevalence of HIV-positive infants born to HIV-positive mothers, 11.4% pooled prevalence in Ethiopia [19] using the assumption of 95% confidence level (Z= Zα/2=95%=1.96), margin of error (d= 5%=0.05), Then, sample size was determined as follows:

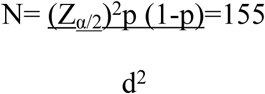

Therefore, the minimum of 155 study participants was selected by consecutive convenience sampling technique.

### Eligibility criteria

All infants born to HIV positive mother attending PMTCT clinic at Shegaw Motta General Hospital were included in this study. While, exposed infants who were critically ill and whose parents were unwilling to give their consent were excluded from the study.

### Data collection and processing

The data were collected by trained a midwifery and principal investigator. Thus, socio-demographic and associated factors were collected by semi-structured pre-tested Amharic version questionnaire using a face-to-face interview with parents of study participants directly after obtaining consents. At each data collection spot, sufficient explanation about the aim of the research was given to the parents before conducting the interview.

### Blood specimen collection, transportation and storage

A minimum of 100µlwhole blood specimen was collected using ethylene diamine tetra acetylene (EDTA) test tube as per the manufacturer’s instruction at heal or toe site of the infants who born to HV positive mother [20]. The collected whole blood was transported immediately from PMTCT clinic to Shegaw Motta General Hospital, Microbiology laboratory for Examination. Due to the Cepheid was busy by which GeneXpert MTB/RIF tests were done and the sample was not done immediately, the samples were stored at 2-8°c, 15-30°c and 31-35°c for up to 72, 24 and 8 hours, respectively [21]. Additionally, 5ml of whole blood was collected from mother with EDTA K3 plasma separating tube (PPT),wait for 4–12hoursand then centrifuged at 3000 rpm for 3minute to separate the 1-2ml plasma from red blood cells. Such separated plasma was stored at 2-8^oC^for a week due to not done immediately. After then, it was transported to Debre Markos Comprehensive specialized hospital (DMCSH), Molecular biology laboratory at 2-8^oC^ with triple packaging system to determine maternal viral load.

### Laboratory testing

#### Early Infant diagnosis (EID) by GeneXpert HIV-1 Qual Assay

In the newly updated algorithm, samples that are nonreactive or indeterminate in the differentiation assay are to be tested with an HIV-1 nucleic acid amplification (NAAT) test for resolution. Xpert HIV-1 Qual assay is a new NAAT assay approved for the identification of HIV infection in whole blood [21].

GeneXpert HIV-1Qual assays were conducted according to manufacturer’s recommendations. EID could be done by the GeneXpert HIV-1 Qual Assay using GeneXpert system machine (Cepheid)[22] by trained laboratory personnel during the study period. The GeneXpert^®^ HIV-1 Qual is a molecular cartridge-based assay that detects total nucleic acid (DNA and RNA) and provides a qualitative result (HIV detectable or undetectable) [23]. As such, the GeneXpert HIV Qual cartridge was labeled with the identification collected blood sample. Then after it was opened and 750 µl of sample reagent was transferred in using provided /inserted pipette in the kit. Mix the whole blood as well by inverting the EDTA tube containing such blood at least seven times. About 100µl whole blood was transferred immediately using provided pipette in the kit or calibrated automatic pipette in to same sample chamber of GeneXpert HIV Qual cartridge soon after the lid was firmly closed. Finally, the prepared GeneXpert HIV Qual cartridge was run by starting the test on GeneXpert machine and interpreted the result output as “HIV-1 detected” or “HIV-1not detected” after 90 minutes [24].

#### Cluster of differentiation at 4 (CD4) Count

Maternal CD4 was counted using FACS Presto Machine after incubating one drop (40µl) out of collected 5ml maternal blood for viral load samples on CD4 cartridge for 18 Minute. Then, assess its association with HIV infection among exposed infants.

#### Maternal Viral Load determination

5ml of whole blood was collected from mother with EDTA K3 plasma separating tube (PPT), wait for 4–12hoursand then centrifuged at 3000 rpm for 3minute to separate the 1-2ml plasma from red blood cells. Such separated plasma was stored at 2-8^oC^for a week due to not done immediately. After then, it was transported to Debre Markos Comprehensive specialized hospital, Molecular biology laboratory at 2-8^oC^ with triple packaging system to determine maternal viral load. Finally, the viral load level was determined by PCR technique(Roche Diagnostics GmbH, Mannheim, Germany), the high or low viral load results were recorded and assessed its association with HIV infection to infants.

### Quality control

The structured questionnaires were prepared in English and translated into Amharic language and then back translated to English to check inconsistencies of meaning of words. About 9 (5%) of structured questionnaire was pretested in Motta health center and training was also provided to one BSc midwifery how to collect the socio-demographic and clinical data. The expired date on the GeneXpert HIV-1Qual cartridges was cheeked and the GeneXpert system machine (Cepheid) was calibrated annual regularly. Quality control for CD4 was also done and printed out daily soon after start up the FACS Presto CD4 Machine and it was cross checked with its standard reference ranges.

### Data analysis

Data was entered by EpiData version 3.1 and data analysis was performed using statistical package for social sciences (SPSS) version 20. The prevalence of HIV-1 was determined by descriptive statistics. Multivariable logistic regression was done by entering the variables with p < 0.2 in the bivariable logistic regression to identify the factors associated with HIV-1 infection among exposed infants by considering the *P* value < 0.05 as a statistically significant association.

### Ethical Consideration

Ethical clearance and permission letter were obtained from Shegaw Motta General Hospital administrative office with reference number (SMGH 534/94/72). Additionally, written consent was obtained from a parent and/or legal guardian of study participants in accordance with the Declaration of Helsinki. Furthermore, the participation of study participants was entirely volunteer based on parents and/or legal guardian and their confidentiality was kept by coding rather than naming for identification. Finally, all HIV positive infants were linked to ART clinic at this hospital for further management.

### Operational definitions

#### GeneXpert HIV-1 Qual assays

GeneXpert® Instrument Systems, is a qualitative in vitro diagnostic test designed to detect human immunodeficiency virus Type 1 (HIV-1) total nucleic acids using human venous whole blood and capillary from individuals suspected of HIV-1 infection for infant.

### Infant

infants or babies whose age less than 12 months and cannot produce their own serological detectable antibodies against HIV.

### HIV1 exposed infants

The infants who born from HIV positive mother or women.

## Results

### Socio-demographic characteristics of study participants

A total of 155 infants were recruited to the current study from which almost more than half of infants 79(50.9%) were females and the rest males. Furthermore, the majority of 87(56.1%), 87(56.1%), 123 (79.4%), 138(89.0%) and 148(95.5%) infants were born from mothers who could not able to write and read, urban residence, self-employed, ages range from 25-34years and married mother, respectively. About 90 (58.1%) infants were less than 6 months in their age and the rest 65 (41.9%) was 6-12months in the current study (Table: 1).

**Table 1:** Socio-demographic characteristics of study participants at Shegaw Motta General Hospital, 2023.

| Variable | Categories | Frequency | Percent (%) |
| --- | --- | --- | --- |
| Gender of infant | Male | 76 | 49.1 |
|  | Female | 79 | 50.9 |
| Residence | Urban | 87 | 56.1 |
|  | Rural | 68 | 43.9 |
| Maternal education | Unable to read and write | 87 | 56.1 |
|  | Primary & above | 68 | 43.9 |
| Maternal age (in years) | < 25 | 6 | 3.9 |
|  | 25–34 | 138 | 89.0 |
|  | ≥35 | 11 | 7.1 |
| Maternal marital status | Not married | 7 | 4.5 |
|  | Married | 148 | 95.5 |
| Maternal occupation | Housewife | 24 | 15.4 |
|  | Government employee | 8 | 5.2 |
|  | Self-employed | 123 | 79.4 |
| Infant age (in months) | <6 months | 90 | 58.1 |
|  | 6- 12months | 65 | 41.9 |

### Prevalence HIV 1 among exposed infants

One hundred fifty-seven HIV-exposed infants were tested for HIV infection by GenXpert HIV1 Qual assay (EID). This study revealed that the overall prevalence of HIV1 infection among exposed infants was 6 (3.87%) with 95% CI; 2.9 - 8.2 in the current study (Fig 1).

**Figure1:**
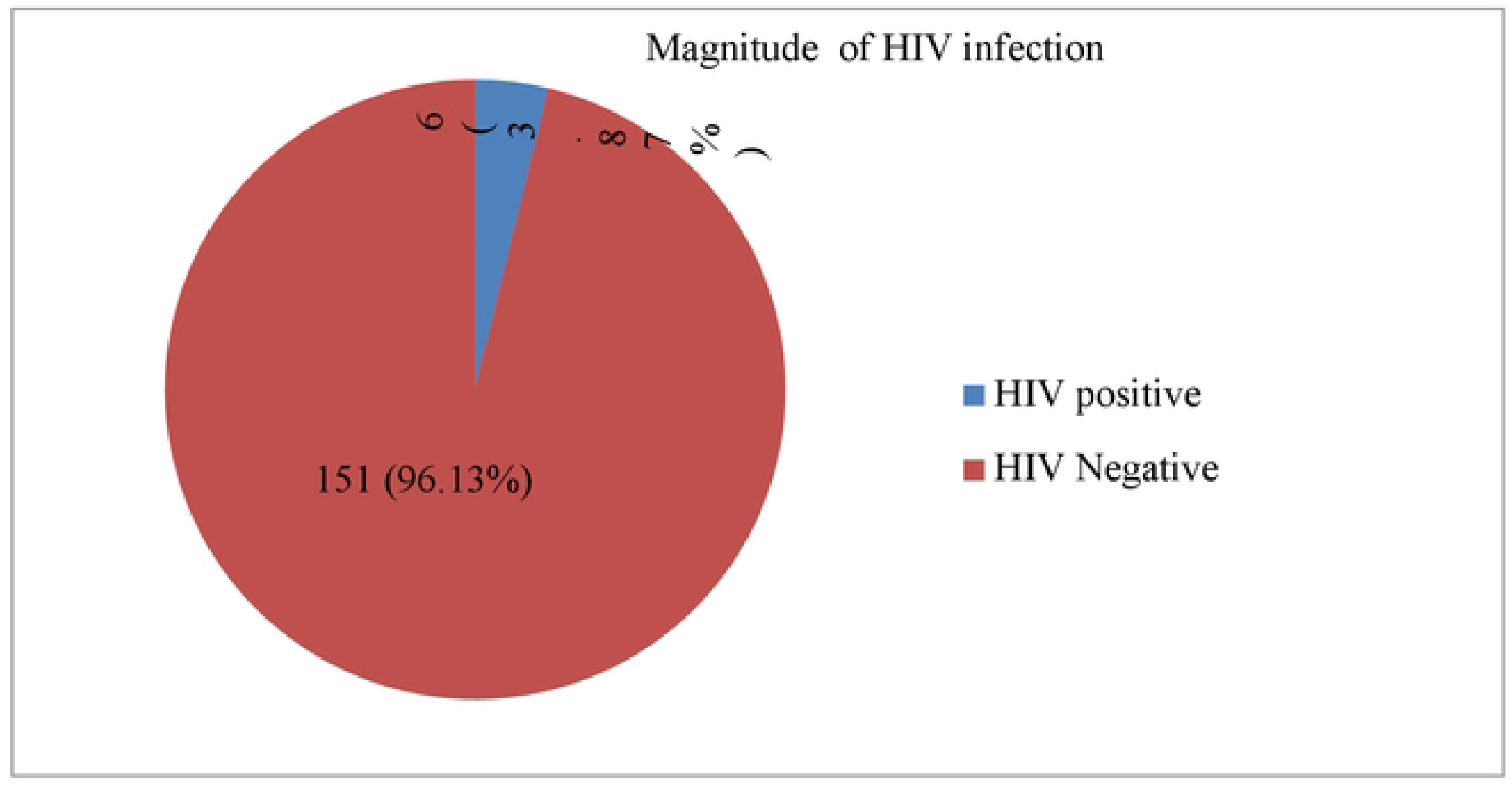
EID positive and negative for HIV1 among exposed infants at Shegaw Motta General Hospital, 2023

### Factors Associated with HIV1 infection among exposed infants

According to bivariable analysis, residence, maternal education level, antenatal care (ANC) follow up, maternal Antiretroviral therapy (ART) enrolment, place of delivery, infant’s age at enrollment, attendant of delivery, maternal Antiretroviral (ARV) prophylaxis, infant nevirapine (NVP) prophylaxis, maternal CD4+ (cell/mm3) and maternal viral load (p-value < 0.2) were entered to multivariable logistic regression analysis. Regarding to multivariable logistic regression analysis, pregnant women had not ANC follow up (AOR=7.281, 95% CI: 2.53-20.96: *P* = 0.001), home delivery (AOR = 3.239, 95% CI: (1.75-9.19, *P*= 0.001), maternal not received ARV prophylaxis (AOR = 9.213, 95% CI: 2.95-10.11, *P* = 0.001), infant not intake NVP prophylaxis (AOR = 2.560, 95% CI: 1.98-10.24, *P* = 0.007) and maternal high viral load (>1000 copies) (AOR = 5.120, 95% CI: 2.75-11.18, *P* = 0.004) during pregnancy were the identified factors significantly associated with HIV infection among infants born to HIV positive mothers. As the result, infants born to HIV positive mother who had not ANC follow up, home delivery, maternal not received ARV prophylaxis, infant not intake NVP prophylaxis and maternal high viral load (>1000 copies) were 7.281, 3.239, 9.213, 2.560 and 5.120 times more likely to be infected by HIV in this study, respectively (Table 2).

**Table 2:** Bivariable and multivariable logistic regression analysis of factors associated with HIV1 infection among exposed infants at Shegaw Motta General Hospital Town, Ethiopia, 2023.

| Variables | Categories | HIV1 | | COR(95%CI) $P$ value | AOR(95%CI) | $P$ value |
| --- | --- | --- | --- | --- | --- | --- |
|  |  | Positive<br>N (%) | Negative<br>N(%) |  |  |  |
| Gender of infant | Male | 2(2.6) | 74(97.4) | 1 |  |  |
|  | Female | 4(5.4) | 71(94.6) | 2.324(0.69-5.61)0.720 |  |  |
| Infant age (in months) | <6 months | 6 (6.7) | 84 (93.3) | 1.653(0.812-7.921)0.431 |  |  |
|  | 6- 12months | 0(0) | 65 (100) | 1 |  |  |
| Residence | Urban | 4(3.4) | 83(96.6) | 1 | 1 |  |
|  | Rural | 2(2.9) | 66(97.1) | 3.12 (1.45-5.11)0.102* | 2.51(0.69-4.67) | 0.145 |
| Maternal education | Unable read and write | 3(3.4) | 85(96.6) | 1.78(2.14-4.76)0.019* | 3.16(0.43-5.89) | 0.205 |
|  | Primary & above | 3(4.5) | 64(95.5) | 1 | 1 |  |
| Maternal age (in years) | < 25 | 0 (0) | 6 (100) | 1 |  |  |
|  | 25–34 | 4 (3.0) | 131 (97.0) | 0.729(0.31-1.73)0.473 |  |  |
|  | ≥ 35 | 2 (18.2) | 9 (81.8) | 1.545(0.47-5.12)0.478 |  |  |
| Maternal marital status | Not married | 1(14.3) | 6(85.7) | 1 |  |  |
|  | Married | 5 (3.4) | 143(96.6) | 1.714(0.72-4.11)0.427 |  |  |
| Maternal occupation | Housewife | 2(8.3) | 22 (91.7) | 0.857(0.31-2.43) 0.367 |  |  |
|  | Self-employed | 3 (2.5) | 120 (97.5) | 1.6490.418-3.24) 0.771 |  |  |
|  | Government employee | 1(12.5) | 7(87.5) | 1 |  |  |
| ANC follow up | Yes | 2 (2.7) | 73 (97.3) | 1 | 1 |  |
|  | No | 4 (5.0) | 76 (95.0) | 10.615(4.40-25.61)0.001* | 7.281(2.53-20.96) | 0.001** |
| Maternal ART enrolment | Enrolled | 2 (1.5) | 135 (98.5) | 1 | 1 |  |
|  | Not enrolled | 4 (23.3) | 14 (77.7) | 16.179(4.58-57.22)0.001* | 6.985(0.61-28.91) | 0.195 |
| Place of delivery | Home | 2(3.9) | 49 (96.1) | 4.681(1.78-12.29) 0.002* | 3.239(1.75-9.19) | 0.001** |
|  | Health facility | 4 (3.8) | 100(96.2) | 1 | 1 |  |
| Infant Feeding pattern | EBF | 0(0) | 23(100) | 1 |  |  |
|  | ERF | 2(3.8) | 50(96.2) | 2.857(0.91-4.43) 0.367 |  |  |
|  | MF | 4(5.0) | 76(95.0) | 1.649(0.518-8.24) 0.571 |  |  |
| Infant's age at enrollment | ≤ 6 weeks | 2(2.0) | 100(98.0) | 1 | 1 |  |
|  | >6 weeks | 4 (7.6) | 49 (92.4) | 4.681(1.78-12.29) 0.002* | 5.219(0.65-14.29) | 0.327 |
| Maternal ARV intervention | Before delivery | 1(2.6) | 38(97.4) | 1 |  |  |
|  | During/after delivery | 2(2.2) | 87(97.8) | 1.857 (0.31-6.43) 0.367 |  |  |
|  | No | 3 (11.1) | 24(88.9) | 4.649(0.418-7.24) 0.771 |  |  |
| Attendant of delivery | TBA | 3(2.9) | 99(97.1) | 4.381(1.68-11.29) 0.001* | 4.239(0.75-9.19) | 0.512 |
|  | Skilled delivery | 3 (5.7) | 50 (94.3) | 1 | 1 |  |
| Maternal ARV prophylaxis | Not Received | 4(11.1) | 32(88.9) | 5.418(2.37-12.41) 0.001* | 9.213(2.95-10.11) | 0.001** |
|  | Received | 2(1.7) | 117 (98.3) | 1 | 1 |  |
| Infant NVP prophylaxis | Yes | 2(2.2) | 90(97.8) | 1 | 1 |  |
|  | No | 4(6.4) | 59(93.6) | 4.681(1.78-12.29) 0.002* | 2.560(1.98-10.24) | 0.007** |
| Maternal CD4+ (cell/mm3) | < 200 | 5(31.2) | 11(68.8) | 1 | 1 |  |
|  | ≥ 200 | 1(0.7) | 138 (99.3) | 5.418(2.37-12.41) 0.001* | 9.213(0.95-10.11) | 0.354 |
| Maternal viral load | <1000copies (Low) | 2(1.4) | 139(98.6) | 1 |  |  |
|  | ≥ 1000 copies)(High) | 4 (28.6) | 10(71.4) | 4.681(1.78-12.29) 0.002* | 5.120(2.75-11.18) | 0.004** |

Key: \* = variables entered to multivariate regression (P- value < 0.2), \*\* = statistical significant association ;ANC: Antenatal care;
AOR: Adjusted odd ratio; ART: antiretroviral therapy; ARV: Anti-retroviral; CD4- cluster of differentiation -4 ; CI: confidence
interval ,COR: crude odd ratio: EBF: exclusive breast feeding ; HIV: human immunodeficiency, MF: mixed feeding; NVP:
Nevirapine , TBA: Traditional birth attendant

## Discussion

The prevalence of HIV1 infection on infants born from HIV positive mothers in the current study was 3.87%. This finding was in line with studies conducted in varies part of Ethiopia like in Dessie Town Public Health Facilities 3.8% [25], Pastoralist Health Facilities, South Omo Zone 5.3% [26], East and West Gojjam Zone5.9% [27] and University of Gondar Specialized Hospital 5.5% [28], east Africa 7.68% (29)and Kenya 3.3% [30].

The current prevalence of HIV1 infection was lower than 11.4% pooled prevalence in Ethiopia [19], 9% in Sidama [31], 10.1% in Amhara Region [32] and 9.9% in Bahir Dar city public health facilities [33]. This difference might be due to the variation of ART and PMCT follow up, awareness to HIV, policies, and strategies on HIV control and prevention, methodology and sample size. In contrast, it was slightly higher than 1.5%, 1.6%, 2.7%, and2.1%, compared to the study conducted in France [34], Ukraine [35], Rwanda[36] and Tigray regional state, Northern Ethiopia [37], respectively. This difference might be due to high coverage of PMTCT interventions in developed countries and limited access, lack of awareness, poor quality of service in developing countries including Ethiopia.

Regarding to multivariable logistic regression analysis in the current study, mothers had not ANC follow up(AOR=7.281, 95% CI: 2.53-20.96: *P* = 0.001)was significantly associated with MTCT ofHIV1. Meanwhile, mothers who did not attend ANC follow up were 7.281 times more likely to transmit the virus to their infants than mothers who had ANC visit. This finding was agreed with the study conducted in Rwanda [38], Ethiopia[39], Gondar city health institutions, Northwest Ethiopia[40] and public health facilities in Dessie town [41].Similarly, home delivery (AOR = 3.239, 95% CI: (1.75-9.19, *P*= 0.001)was also a factor significantly associated with

MTCT of HIV1 infection. Thus, infants born at home had a 3.239 times greater chance of contracting the virus than those born in health facilities. This finding was concordant with the study illustrated in East Africa [42], rural Uganda [43], South west Ethiopia [44], Dire Dawa [45], southern Ethiopia [46], Northwest Ethiopia [47], Gondar city health institutions [40],South Gondar zone [48] and Bahir Dar administration(49).This might be due to HIV testing for mothers with unknown HIV status who delivered at health facility and immediate ARV intervention for mothers and their infants if the test was positive. In addition, safe delivery practice and appropriate post-natal care during health facility delivery might support this significant association.

On the other hand, absence of maternal intake ARV prophylaxis(AOR = 9.213, 95% CI: 2.95-10.11, *P* = 0.001) was 9.213 times more likely to born HIV positive infants compared to those received ARV prophylaxis. A finding was in line with studies conducted in Vietnam[50],East Africa [42],Uganda[43], Northwest Ethiopia[47],Ethiopia such as Mekele city [51], health facilities of North Wollo Zone [52], Bahir Dar administration[49]. This might be as a result of maternal ARV drug intake causing the reduction of maternal viral load and reduced risk of viral transmission to their infants. Furthermore, infants not intake NVP prophylaxis (AOR = 2.560, 95% CI: 1.98-10.24, *P* = 0.007)were2.560times more likely to be HIV positive as compared to infants received NVP prophylaxis at birth according to this study. Such findings were agreed with the previous studies reported in Brazil [53], Uganda (54), Dire Dawa [45], southern Ethiopia [55] and Ethiopian Public Health Institute [56, 57]. This might be due to the viral suppression effect of NVP, which is a non-nucleoside reverse transcriptase inhibitor, by binding to reverse transcriptase, thereby blocking RNA and DNA dependent DNA polymerase actions including HIV replication.

Finally, maternal high viral load (>1000 copies) (AOR = 5.120, 95% CI: 2.75-11.18, *P* = 0.004) during pregnancy were the factors significantly associated with HIV infection among exposed infants by which infants from high maternal viral load were 5.120times more likely to be infected by HIV than infants born from low maternal viral load. The findings were concordant with a study carried out in India [58], Philadelphia [59], Uganda (54), Rwanda [38], Gaza Province—Mozambique (60), northeast South Africa [61]. This might be due to high viral load concentration could compromise the maternal immune system and leads to vertical transmission of HIV to infants.

## Conclusion

The prevalence of HIV1 infection among exposed infants born to HIV positive mothers was still high (3.87%). Pregnant women had not ANC visits, home delivery, Absence of ARV prophylaxis, infants not intake NVP prophylaxis, and maternal high viral load increases HIV infection among exposed infants. Therefore, health facilities should strictly strengthen the PMTCT service by providing maternal ARV prophylaxis, infant NVP prophylaxis, promote ANC service, early screening maternal viral load and scale up skilled delivery to eliminate HIV infection among exposed infants.

## Data Availability

* correspondence author

## Abbreviations

AOR: adjusted odd ratio
ART: Antiretroviral therapy
ARV: antiretroviral
ANC: Antenatal Care
COR: Crude odd ratio
DNA: Deoxy-ribose nucleicacid
EDTA: ethylenediamine tetra acetylene
EID: early infant diagnosis
HIV: Human immunodeficiency virus
NAAT: nucleic acid amplification
NVP: nevirapine
PCR: polymerase chain reaction
PMTCT: prevention of mother-to child transmission,
MTCT: mother to child transmission
RNA: ribose nucleic acid

## Acknowledgments

The authors would like to thank administration of Shegaw Motta General Hospital for providing a permission to conduct this study. By the next, we would like to acknowledge Molecular biology laboratory of Debre Markos comprehensive specialized hospital (DMCSH) including its laboratory staffs that did the viral load of referred plasma sample. We would also like to thank all laboratory staffs in this hospital, study participants, and data collectors for their unreserved efforts and willingness to participate in this study.

## Authors Contribution

**Conceptualization**: Destaw Kebede Nigusie, Fantahun Getaneh Damitew,

**Data curation**: Fantahun Getaneh Damitew, Kirubel Endalamaw Melsew

**Formal analysis**: Destaw Kebede Nigusie, Melsew, Girma Zerefaw, Abebe Fenta Nigusie :

**Investigation:** Destaw Kebede Nigusie, Fantahun Getaneh Damitew, Kirubel Endalamaw Melsew, Girma Zerefaw, Abebe Fenta Nigusie

**Methodology**: Kirubel Endalamaw Melsew, Girma Zerefaw, Abebe Fenta Nigusie

**Resources:** Destaw Kebede Nigusie and Fantahun Getaneh Damitew

**Supervision:**, Girma Zerefaw Abay, and Abebe Fenta Nigusie

**Visualization:** Fantahun Getaneh Damitew, Kirubel Endalamaw

**Writing – Original Draft Preparation:** Destaw Kebede Nigusie, Fantahun Getaneh Damitew,

**Writing – Review & Editing:** Girma Zerefaw Abay, and Abebe Fenta Nigusie

